# GLP-1 Receptor Agonist Initiation and Anti-VEGF Treatment Frequency in Diabetic Macular Edema: an IRIS^®^ Registry Cohort Study

**DOI:** 10.64898/2026.08.29.26361426

**Authors:** Preeti Nagalamadaka, Connor Ross, Joshua B. Gilbert, Hannah Stillman, Sophia Y. Ghauri, Sophia M. Dutton, William Kearney, Josephine H. Li, Aaron Leong, Rishi P. Singh, Magdalena G. Krzystolik

## Abstract

**Purpose:** To evaluate whether initiation of GLP-1 receptor agonists (GLP-1RAs) is associated with anti-VEGF treatment burden in type 2 diabetes patients with diabetic macular edema (DME) in the IRIS^®^ Registry (Intelligent Research in Sight).

**Methods:** Incident GLP-1RA initiators were matched 1:1 with controls via Mahalanobis distance matching (9,896 pairs; N=19,792) on sociodemographics, DME risk factors, and factors influencing GLP-1RA prescription including hypertension, obesity, chronic kidney disease. A longitudinal mixed-effects event-study model evaluated monthly anti-VEGF injection frequency over a 36-month window (12 months before through 24 months after initiation), adjusting for DME duration. Visual acuity (VA) and central subfield thickness (CST) were secondary outcomes.

**Results:** Following GLP-1RA initiation, anti-VEGF injection trajectories did not significantly differ between the matched GLP-1RA and control cohorts (interaction coefficients −0.18 to 1.59, P>0.05). Likewise, no differences in VA were observed between cohorts (−0.05 to 0.04 logMAR, P>0.05) or CST (−14.12 to 33.58 µm, P>0.05).

**Conclusion:** In these matched cohorts, GLP-1RA initiation was not associated with the trajectory of anti-VEGF use or changes in VA or CST.

**Précis:** We used the American Academy of Ophthalmology IRIS^®^ Registry (Intelligent Research in Sight) to identify patients with DME. In 19,792 matched patients, there was no significant reduction in injection frequency post GLP1-RA initiation and no significant change in VA or CST.

## Introduction

Diabetic macular edema (DME) is a complication of diabetic retinopathy (DR) in which hyperglycemia-induced damage to retinal vasculature leads to fluid leakage into the macula, causing retinal thickening and swelling that can result in vision loss^1^. DME affects one in 15 people with diabetes resulting in more than 20 million cases worldwide^2^. The extent of macular swelling and its functional impact are measured via central subfield thickness (CST) and visual acuity (VA).

Intravitreal anti-vascular endothelial growth factor (anti-VEGF) therapy has transformed the management of DME. With the introduction of anti-VEGF agents, treatment of DME has been revolutionized, substantially limiting the role of laser photocoagulation. Both the European Society of Retina Specialists (EURETINA) and the American Academy of Ophthalmology (Academy) recommend anti-VEGF agents as the preferred first-line pharmacotherapy for DME, with intravitreal corticosteroids reserved as a second-line option due to side effects including secondary glaucoma and cataract formation^3^. Anti-VEGF agents such as ranibizumab, bevacizumab, and aflibercept restore blood-retinal barrier integrity, resolve macular fluid, and improve vision in most patients by binding and neutralizing soluble VEGF. The frequency of injections reflects both the underlying disease activity and clinician-driven treatment regimens, such as fixed interval protocols or treat-and-extend approaches.

Treatment response is incomplete in a meaningful subset of patients due to poor glycemic control, renal impairment, and diabetes duration over 15 years^4,5^. The burden of frequent intravitreal injections, often monthly or bimonthly, represents a significant challenge for both those patients and healthcare systems. Many patients with DME do not achieve complete fluid resolution despite multiple anti-VEGF injections, underscoring the need to understand factors that may modify treatment requirements. One such factor may be overall glycemic control. Glucagon-like peptide-1 receptor agonists (GLP-1RAs) are a class of medications used to treat type 2 diabetes and obesity that mimic the endogenous incretin hormone GLP-1, improving glycemic control through multiple mechanisms, including enhanced glucose-dependent insulin secretion, suppression of glucagon secretion, delayed gastric emptying, and increased satiety. This leads to significant weight loss and lowering of blood glucose levels, proving highly therapeutic for patients with diabetes^6^. In 2024, 26.5% of patients with diabetes were being treated with a GLP-1RA^7^.

Previous work has attempted to address whether diabetic treatment with a GLP-1RA also reduces DR and DME. Early cardiovascular outcomes trials, most notably SUSTAIN-6, raised concern for worsening DR complications among semaglutide-treated patients, prompting concern that GLP-1RA therapy may accelerate retinal microvascular disease^8^. However, this finding has generally been interpreted within the context of rapid glycemic improvement and the well-described phenomenon of early worsening, rather than a direct retinotoxic effect^9^. There have been later studies showing conflicting results of the effect of GLP-1RA on diabetic eye disease. Some studies have reported decreased likelihood of developing DR or DME when patients are treated with GLP-1RA^10,11^, while some reported equal^12–18^ or even increased^19–21^ likelihood. It is unclear if the decrease in blood sugar halts or even reverses inflammation, or whether it triggers a sudden increase in inflammation, worsening DR. Critically, prior studies have largely focused on incident DR or DME development rather than on disease progression in patients already established on anti-VEGF treatment. To address this knowledge gap, we evaluated the association of GLP-1RA initiation with anti-VEGF injection frequency, VA, and CST among patients with existing DME in a large ophthalmology registry.

## Methods

This study used a retrospective matched cohort design, controlling for medications and comorbidities, to evaluate the association between GLP1-RA use and changes in anti-VEGF injection frequency in patients with DME (due to Type 2 diabetes).

### Data Source

The American Academy of Ophthalmology IRIS^®^ Registry (Intelligent Research in Sight) is a centralized data repository and reporting tool that can be used for research purposes. The registry captures longitudinal clinical data from 50 million patients per year and includes diagnoses via International Classification of Disease 10 (ICD-10) codes, treatments via procedures with Current Procedural Terminology (CPT) codes, pharmacy fills via RXCUI ingredient codes, visual outcomes, and patient demographics. We used the IRIS Registry specifically for this study because it offered an avenue to study the progression of DME in relation to GLP1-RA use in a large-scale nationally representative sample. The analytic dataset was structured with one record per month for each patient (one eye randomly selected). The index event was the date of DME diagnosis.

The IRIS Registry is a centralized data repository and reporting tool that can be used for research purposes. This does not constitute human subject research because data in the IRIS Registry is de-identified and the investigator does not have access to study identifiers. Therefore, institutional board review and informed consent are not required. This study adheres to the Declaration of Helsinki.

### Cohort Selection and Matching

We identified adult patients (age ≥18 at diagnosis) with a documented DME diagnosis who received an anti-VEGF injection at or within four weeks of their DME diagnosis date (Supplemental Table 1). To be eligible as a GLP-1RA initiator (treated cohort), patients must have initiated a GLP-1RA after their DME diagnosis date with no prior GLP-1RA prescription recorded in the IRIS Registry. Patients were excluded if they had a history of retinal diseases that could have affected the macula (Supplemental Table 2). We randomly selected one eye per patient if both eyes were eligible for inclusion.

To reduce confounding by observed characteristics, we applied Mahalanobis Distance Matching (MDM) to construct 1:1 matched pairs of GLP-1RA initiators and non-initiators^22^. Matching covariates included DME diagnosis date, last observed date in the IRIS Registry, age at DME onset, sex, race and ethnicity, median household income category, neighborhood high school graduation rate, urban/rural classification, tobacco usage, insulin usage, severe obesity diagnosis, antihyperglycemic use, severity of proliferative diabetic retinopathy, history of vitreomacular adhesion and baseline mean visual acuity (mean logMAR visual acuity 12 months prior to DME diagnosis). We also matched on factors that could influence a patient’s use of a GLP1-RA including history of acute pancreatitis, coronary artery disease, chronic kidney disease, gastroparesis and other specified conditions that were available in the dataset (Supplemental Table 3). Notably, we were unable to match for HbA1c or body mass index; we discuss the implications of this limitation further below. We matched on DME diagnosis date and last observed date to ensure each GLP-1RA user and their matched control had comparable observation windows, enabling valid longitudinal comparisons between matched pairs. Balance across matched cohorts was assessed by examining standardized mean differences (SMDs) and the distribution of baseline characteristics between cohorts. We also used the Wilcoxon rank-sum test for continuous variables and the Pearson chi-square test for categorical variables. The final matched sample consisted of 9,896 patients (one eye per patient) per cohort (N = 19,792 total).

### Statistical Analysis

For each patient, in each month, we recorded whether they received an anti-VEGF injection during that month or the one before it, with 1 indicating yes and 0 indicating no. By using this dichotomous indicator as our outcome variable, we obtain the probability (multiplied by 100 for percentage) that a patient receives an injection in any given month. In our longitudinal model, this allows us to test how anti-VEGF injection probabilities may evolve differently among matched patients who do or do not initiate GLP-1RA treatment. Since patients started GLP-1RAs at different times relative to DME diagnosis, we coded the data to align all patients at a common starting point. For GLP-1RA patients, “month 0” represents the month the patient began a GLP-1RA, “month −1” represents the month prior to initiation, “month +1” represents the month post initiation, and so forth. For matched control patients who do not initiate a GLP-1RA, we set month “0” to be the number of months from DME diagnosis that the treated patient initiated GLP-1RA. For example, if a treated patient initiates GLP-1RA five months post DME diagnosis, we set the fifth month from DME diagnosis in the matched control patient to serve as “month 0” for comparison. We then tracked each patient for up to 12 months before and 24 months after that starting point. Control patients were required to remain observed in the registry through the assigned month 0 to contribute follow-up data, so this alignment strategy carries a potential for selection bias if attrition from the registry was differentially associated with the outcome. This is not fully addressed by matching on last observation and initial diagnosis date.

We used a linear mixed effects model to estimate the effect of GLP-1RA initiation on anti-VEGF injection probability over time. Covariates included a treatment group indicator, month fixed effects, and their interaction (yielding month-by-month estimates of the GLP-1RA effect). The model estimated, for each month in the follow-up window, whether GLP-1RA users and controls differed in their trends in the probability of receiving an injection, after adjusting for how long each patient had been diagnosed with DME. Because the same patients were observed repeatedly over time, the model also accounted for the fact that injection patterns tend to be correlated within a person across months by including a random intercept for the patient. Since we include pre-initiation interaction terms (months −12 through −1), the model allows a formal assessment of pre-treatment trend parallelism; the post-initiation interaction terms represent the differential change in anti-VEGF probability attributable to GLP-1RA initiation under the assumption that the trends would have continued in parallel in the absence of treatment^23^. The reference month was −1, which was the month immediately prior to initiation. Secondary outcomes were visual acuity (VA, in logMAR) and central subfield thickness (CST, in µm).

All analyses were conducted in R, version 4.5.1. The statistical significance threshold was set at α = 0.05.

## Results

### Cohort Demographics

The matched GLP-1RA and control cohorts were balanced on most demographic and clinical characteristics, though several variables reached statistical significance by conventional hypothesis testing given the large sample size (N=19,792) (Table 1). We therefore report both p-values and SMDs in Table 1, as SMDs are less sensitive to sample size and better reflect the practical magnitude of between-group differences. SMDs were not large enough to be clinically meaningful. Sex distribution was approximately equal (∼50% female in each group). Racial and ethnic composition was similar: ∼47-48% White, ∼11% Black or African American, ∼13-14% Hispanic or Latino. Most patients resided in urban areas (∼85-86%). Tobacco use and neighborhood-level socioeconomic indicators were comparable between groups (Table 1).

**Table 1.**
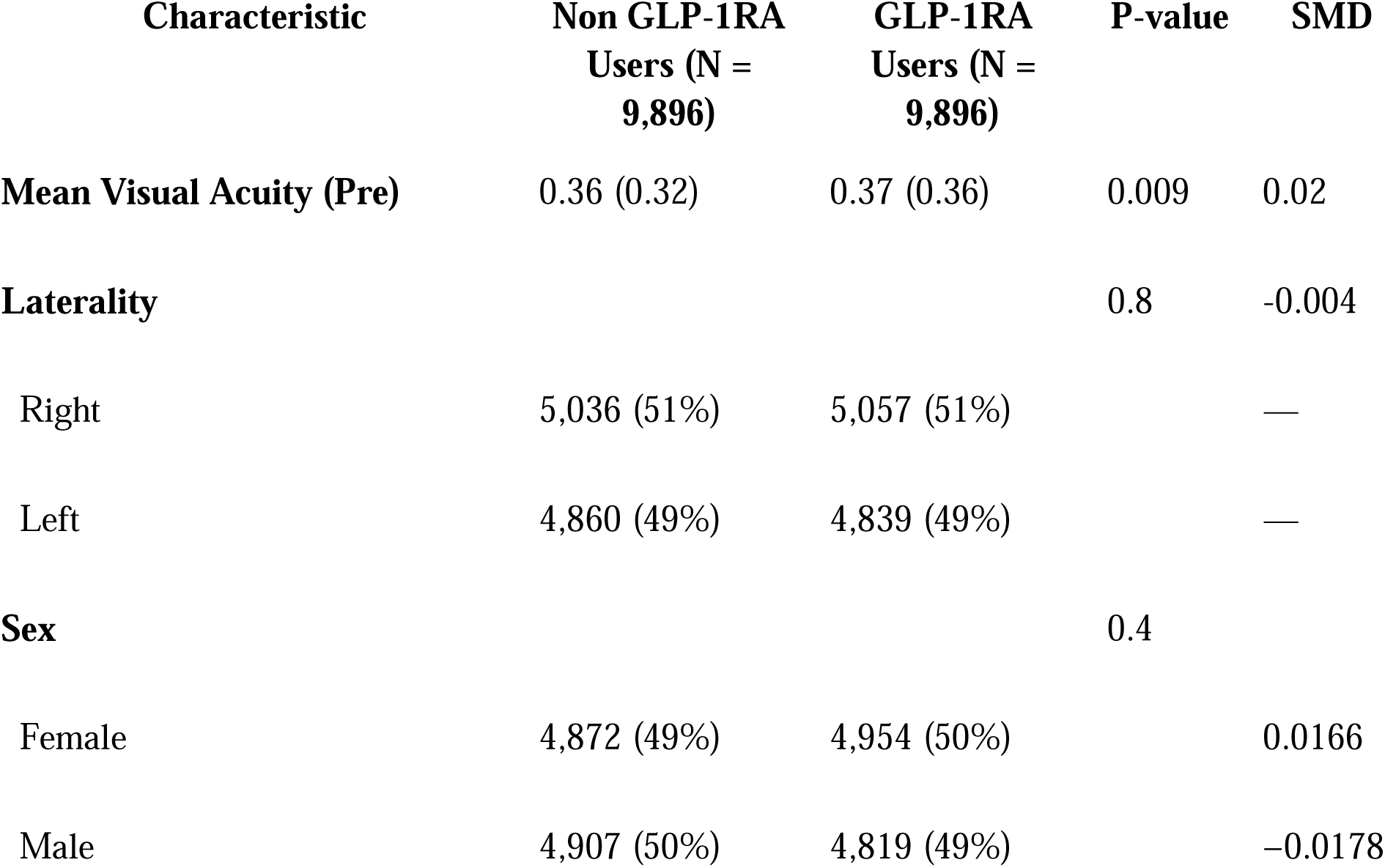
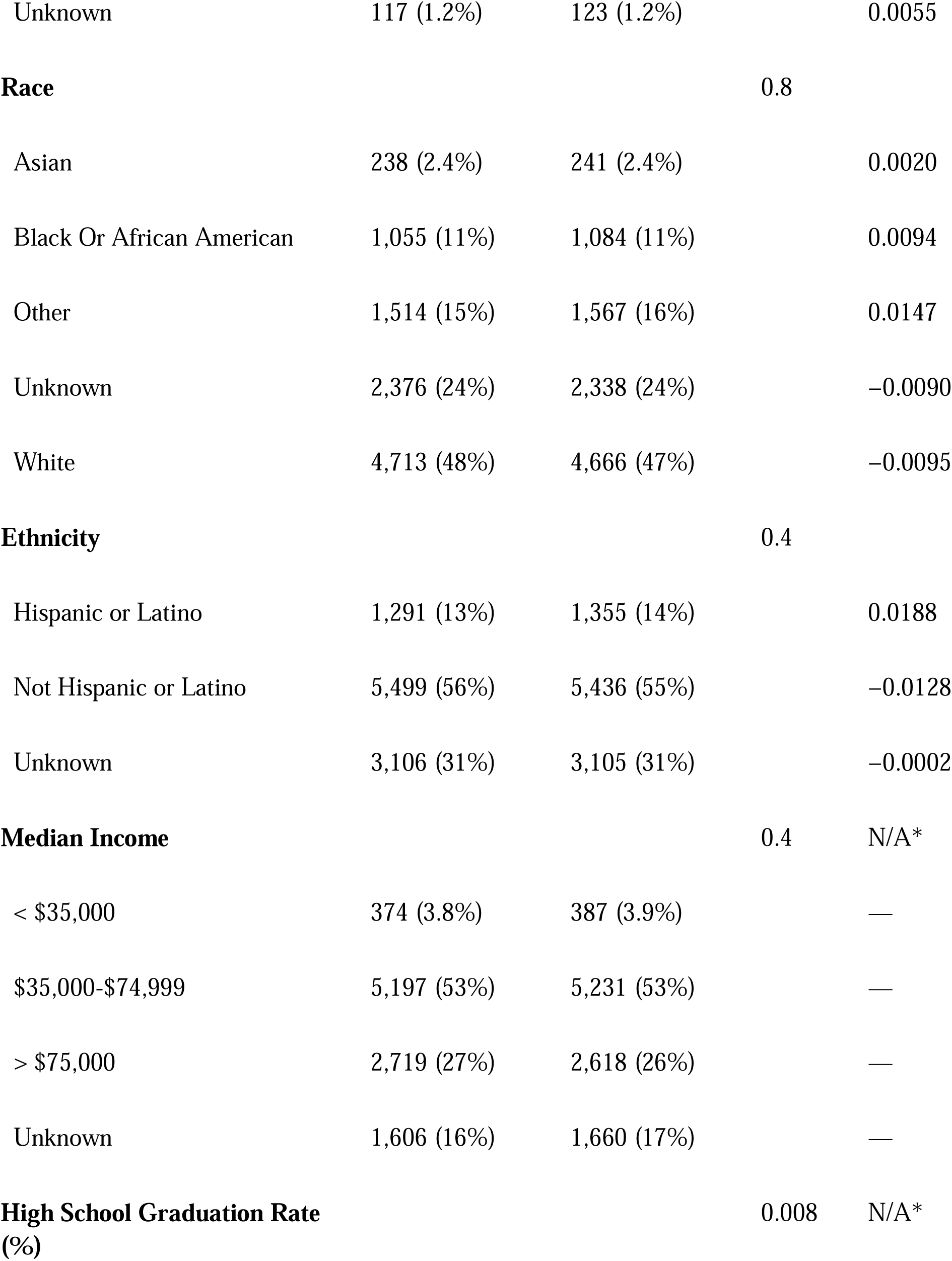
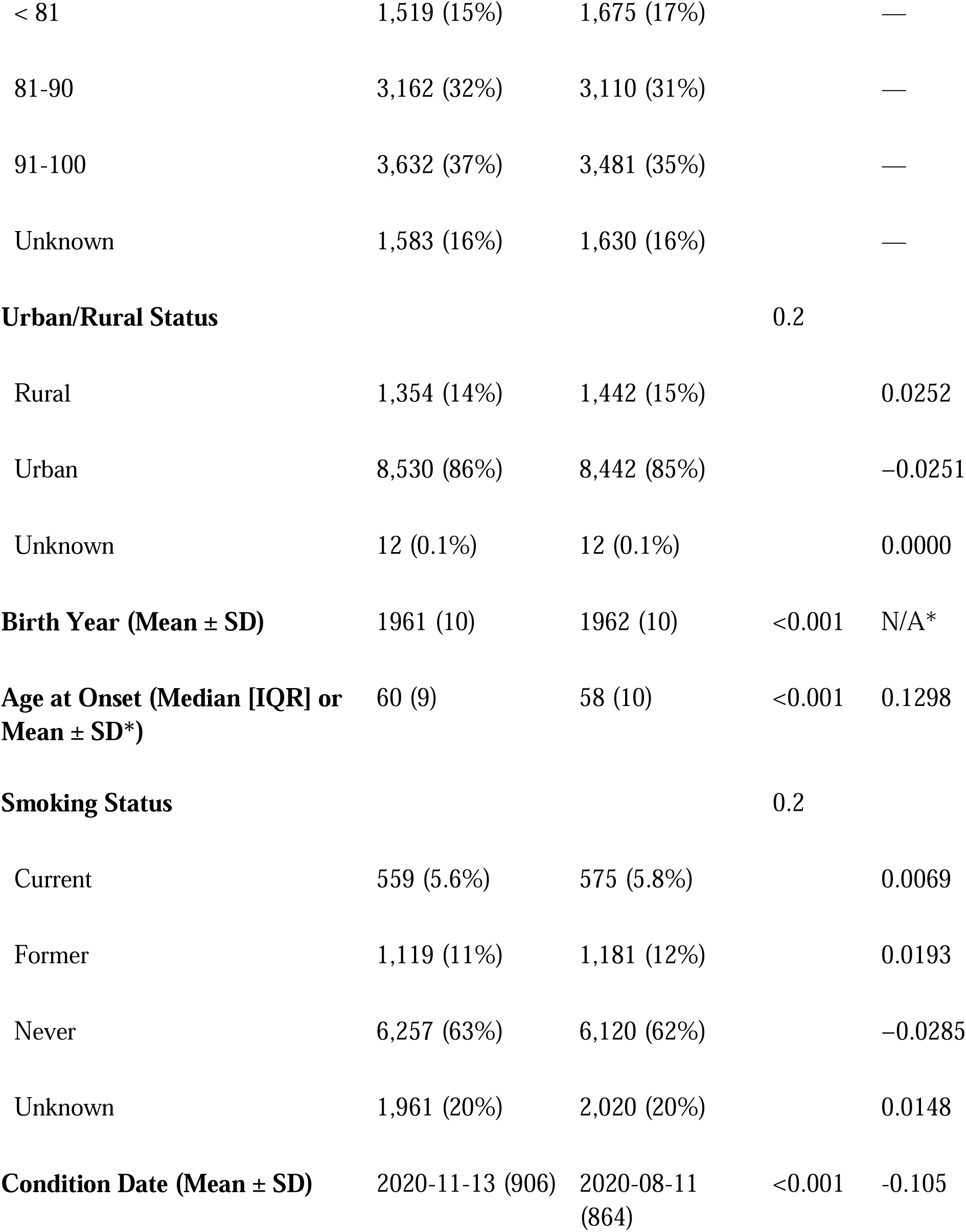
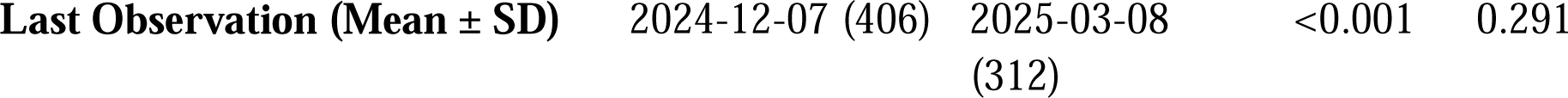
Baseline Demographic and Clinical Characteristics of the Study Population. Values are presented as mean (standard deviation) for normally distributed continuous variables, median (interquartile range) for non-normally distributed continuous variables, and count (percentage) for categorical variables. P-values were calculated to compare differences between the two cohorts using the Wilcoxon rank-sum test for continuous variables and the Pearson chi-square test for categorical variables. Standardized mean difference (SMD) was also calculated.

### Anti-VEGF Probability

The intercept for the control cohort one month before GLP-1RA initiation was approximately 17.1 percentage points, which is about a 17% probability of anti-VEGF injection in a given month in the first year from DME diagnosis. GLP-1RA patients had a lower baseline injection probability overall one month before GLP-1RA initiation (main effect of GLP-1RA cohort: −5.55 percentage points, SE = 0.45, P<0.001) (Figure 1). Both cohorts followed similar overall trajectories, with no evidence of a trend divergence following GLP-1RA initiation (Figure 1).

**Figure 1.**
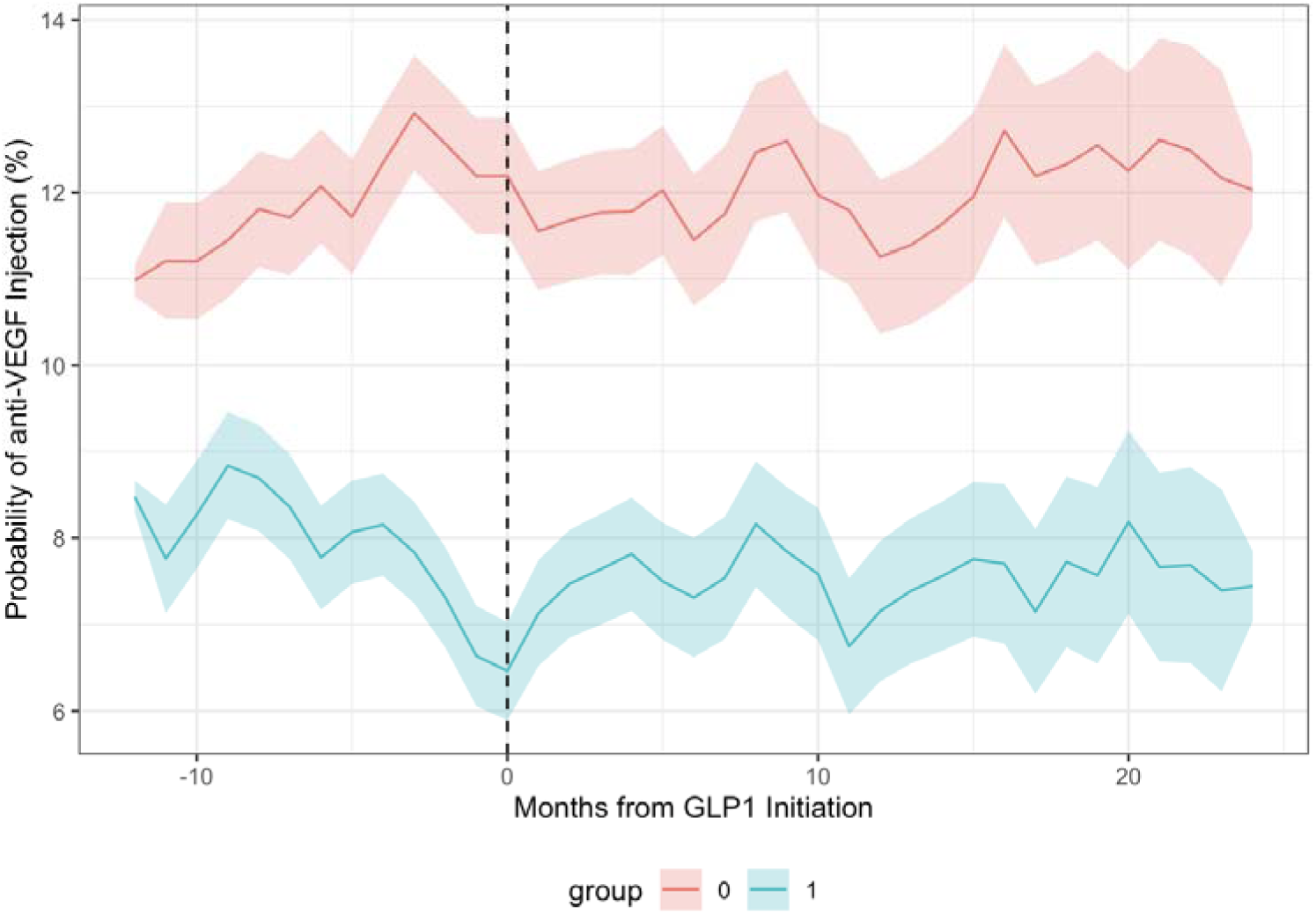
Estimated monthly probability of anti-VEGF injection for GLP-1RA initiators (Group 1) and matched controls (Group 0) for the average patient. The vertical dashed line indicates the month of GLP-1RA initiation in the treated group (Month 0). Shaded areas represent 95% confidence intervals.

This baseline difference indicates that GLP-1RA initiators and controls differed systematically in injection probability prior to initiation, a channeling pattern consistent with GLP-1RA initiation occurring preferentially among patients with somewhat lower baseline treatment intensity. However, because our causal interpretation rests on the parallel pre-trends assumption rather than on equivalence of baseline levels, and because pre-initiation interaction terms (months −12 through −1) showed no evidence of trend divergence, this level difference does not undermine the validity of the difference-in-differences estimate, provided the parallel-trends assumption holds.

Following initiation, the trajectories remained stable and parallel through 24 months of follow-up. The difference-in-trends analysis (Figure 2) confirmed that GLP-1RA initiation did not result in a sustained or progressive reduction or increase in treatment burden; the estimated difference between cohorts remained near zero and lacked a consistent slope over the two-year post-treatment period.

**Figure 2.**
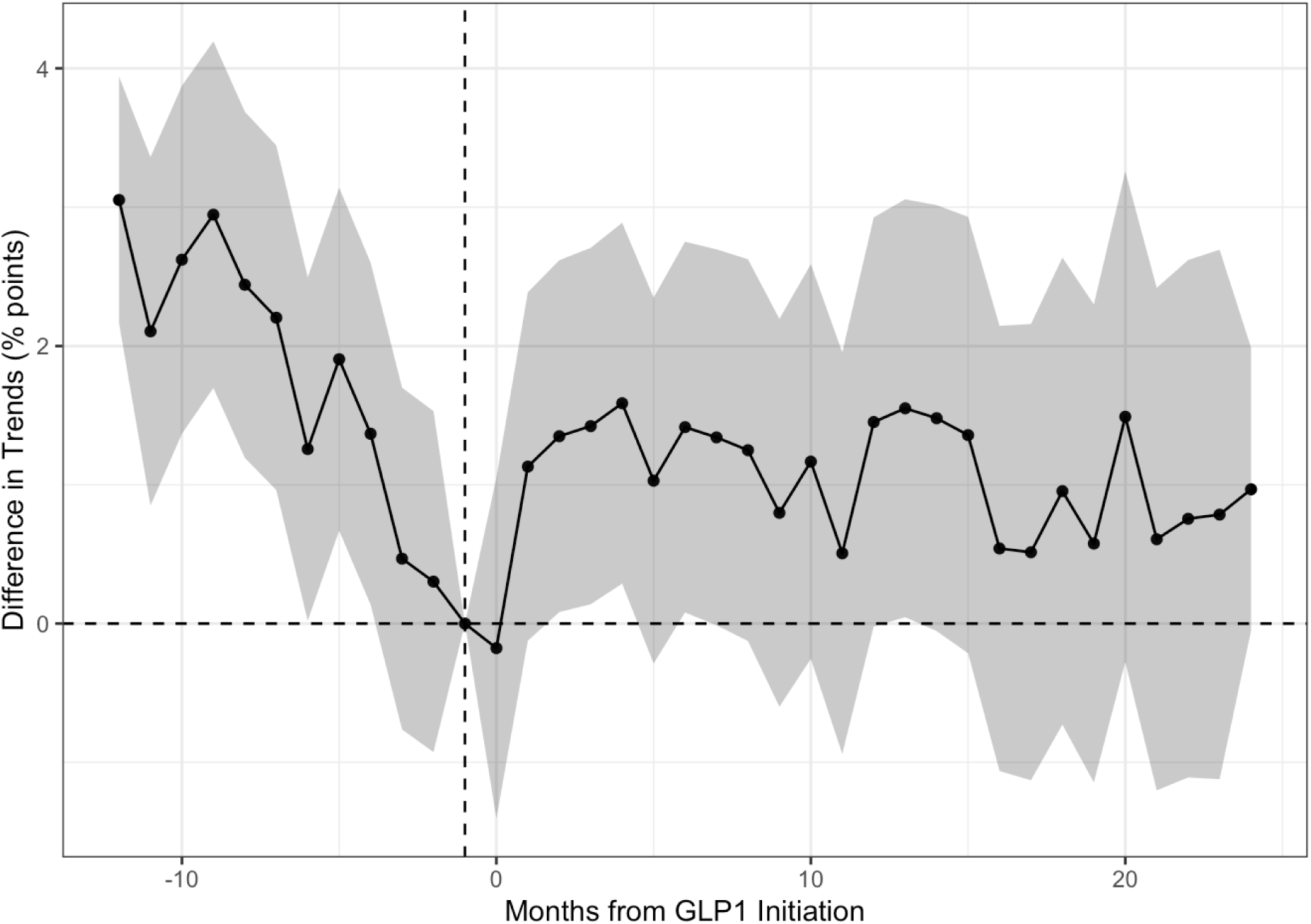
Difference in anti-VEGF injection probability trends between the GLP-1RA cohort and matched controls. Month −1 is fixed to 0 as the reference category.

### Visual Acuity and Central Subfield Thickness

The GLP-1RA cohort showed no significant overall difference in VA or CST trajectory compared to controls. The interaction terms for VA (logMAR) ranged from −0.05 to +0.04 across relative months (Figure 3). CST interaction coefficients ranged from −14.12 to +33.58 µm (Figure 4). No statistically significant or directionally consistent effects of GLP-1RA initiation on VA or CST were detected.

**Figure 3.**
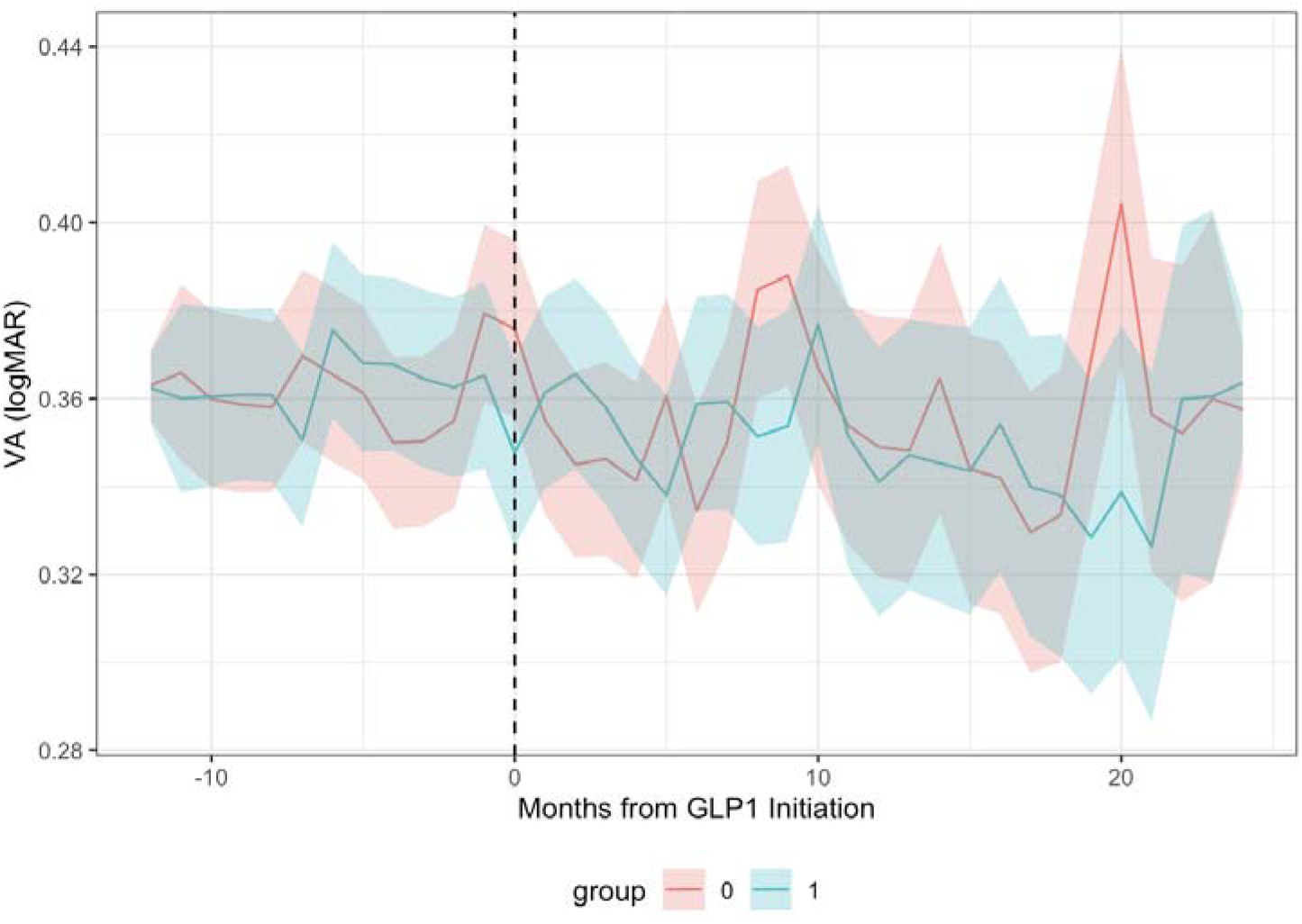
Mean visual acuity (measured in logMAR) for GLP-1RA initiators and matched controls over the 36-month study window. GLP-1RA initiators are Group 1 and matched controls are Group 0.

**Figure 4.**
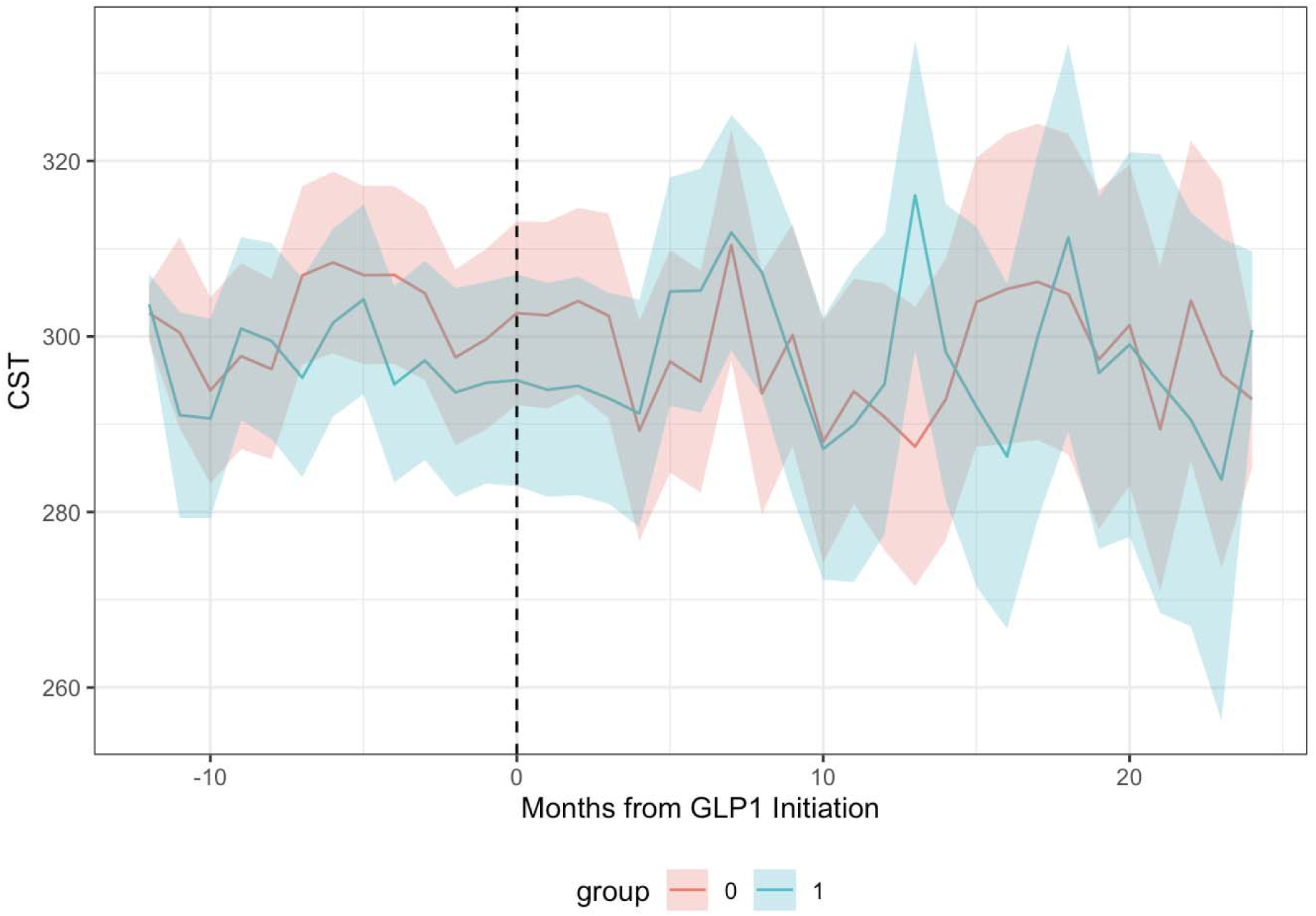
Mean central subfield thickness (measured in µm) for GLP-1RA initiators and matched controls over the 36-month study window. GLP-1RA initiators are Group 1 and matched controls are Group 0.

## Discussion

In this large longitudinal matched-cohort study, evaluating GLP-1RA exposure in patients with DME receiving anti-VEGF therapy, initiation of GLP-1RA treatment was not associated with any significant change in anti-VEGF treatment burden, VA, or CST over 24 months post GLP-1RA initiation. These consistently null findings across all three clinically relevant outcome domains support a clinically neutral association between GLP-1RA initiation and DME outcomes. Collectively, these findings provide reassuring evidence that GLP-1RA therapy does not appear to adversely alter the clinical course of DME in patients established on anti-VEGF treatment and should support greater confidence in its use when clinically indicated for systemic metabolic benefit.

The relationship between GLP-1RA exposure and diabetic retinal disease remains an area of active investigation, with prior literature reporting mixed findings. Some population-based studies suggest reduced rates of incident DR or DME among GLP-1RA users, while others have demonstrated no significant association with DR progression, DME development, or treatment-requiring retinal disease^24–28^. More recent studies have increasingly supported a neutral ophthalmic association. In a large TriNetX analysis, Ramsey et al. found that although GLP-1RA exposure was associated with a modest increase in incidence of DR, it was not associated with progression to proliferative DR or incident DME in eyes with pre-existing DR, and was instead associated with lower rates of vitreous hemorrhage, neovascular glaucoma, blindness, and need for retinal intervention.^26^ Building on these prior studies, we assessed patients already undergoing active anti-VEGF treatment for established DME, which could be practically helpful for retinal specialists. Talebi et al. reported that, after adjustment for longitudinal glycemic control and key confounders, GLP-1RA use was associated with lower risk of DR, DME, and treatment-requiring DR/DME over 10 years.^29^ In contrast, Wai et al. observed a higher relative risk of progression to proliferative DR and DME in GLP-1RA users compared with matched SGLT-2 inhibitor users.^30^ Building on the foundational contributions of Ramsey, Talebi, and Wai, our study extends this body of work by focusing specifically on patients with active DME already receiving anti-VEGF therapy, using injection frequency as a direct and clinically meaningful measure of disease activity that also reflects real-world treatment burden, and additionally tracking VA and CST trajectories, offering a complementary and treatment-relevant perspective.

This study has several limitations. All included eyes were already receiving anti-VEGF therapy, which substantially reduces macular edema activity and may limit the incremental detectable impact of systemic metabolic interventions. Another limitation is the lack of patient-level metabolic measurements within the IRIS Registry. Although we matched on numerous demographic characteristics, cardiometabolic comorbidities, concomitant medications, PDR, and DME duration, we were unable to account for measures of diabetes severity (e.g., baseline and longitudinal HbA1c, diabetes duration, and insulin treatment intensity) and body mass index. As GLP-1RAs are typically initiated in response to worsening metabolic status, residual confounding by indication remains possible. Incomplete adjustment may have attenuated a true association or obscured a modest treatment effect. Because retinal effects of GLP-1RAs are likely mediated through improvements in metabolic control, these unmeasured factors are likely relevant to the interpretation of our findings. Since control patients were required to remain observed in the registry through the assigned month 0 in order to contribute follow-up data, this alignment strategy carries a potential for selection bias if attrition from the registry was differentially associated with the outcome, which may not be fully addressed by matching on last observation and initial diagnosis date. As a retrospective analysis of the IRIS Registry, the study relies on billing and diagnostic coding, which limits clinical phenotyping granularity and precludes full adjustment for key systemic variables, including longitudinal HbA1c trajectories. Residual confounding from unmeasured clinical variables therefore remains possible despite multivariable matching. Our study captures GLP-1RA initiation but lacks data on actual patient adherence and continuation of GLP-1RA. The 24-month post-initiation follow-up window, while substantial for a registry-based study, remains short relative to the chronic, progressive natural history of diabetic microvascular disease, and it is possible that a true effect of GLP-1RA therapy on DME course would only become detectable over a longer observation period. Future studies incorporating longitudinal glycemic control, medication adherence, and cumulative GLP-1RA exposure may further clarify whether specific patient subgroups derive retinal benefit.

Our findings suggest that initiation of a GLP-1 receptor agonist does not meaningfully alter the clinical course or treatment burden of DME compared with other established treatment pathways. We recommend that the management of DME should therefore continue to be guided primarily by ocular disease activity and evidence-based use of anti-VEGF therapy, with systemic antidiabetic selection remaining a parallel but independent consideration.

## Conclusion

In this large-scale matched cohort study of 19,792 patients drawn from the IRIS Registry, GLP-1RA initiation was not associated with any significant change in anti-VEGF injection frequency, visual acuity, or central subfield thickness over 24 months of follow-up. Across all three clinically relevant outcome domains, GLP-1RA therapy demonstrated a retinally neutral profile, neither reducing treatment burden nor accelerating anatomical or functional deterioration in patients with established DME receiving anti-VEGF therapy. These findings are meaningful given the ongoing uncertainty in the literature regarding the ophthalmic safety of GLP-1RAs, including early signals of retinopathy worsening from cardiovascular outcomes trials. Our results provide reassuring data that GLP-1RA initiation for systemic metabolic or weight-related indications may be unlikely to adversely affect the clinical course of DME or require modification of existing retinal treatment standards. DME management should continue to be guided by intraocular disease activity and anti-VEGF treatment response, with systemic antidiabetic therapy selection remaining an independently determined clinical decision.

## Data Availability

All data referenced in this manuscript are derived from the American Academy of Ophthalmology IRIS Registry. The Registry is a de‑identified, HIPAA‑compliant clinical database comprising tens of millions of patient encounters and accessible to researchers via AAO-approved pathways.

## References

1. Skarbez K, Priestley Y, Hoepf M, Koevary SB. Comprehensive Review of the Effects of Diabetes on Ocular Health. Expert Rev Ophthalmol. 2010;5(4):557–577. doi:10.1586/eop.10.44

2. Tan GS, Cheung N, Simó R, Cheung GCM, Wong TY. Diabetic macular oedema. Lancet Diabetes Endocrinol. 2017;5(2):143–155. doi:10.1016/S2213-8587(16)30052-3

3. Lim JI, Kim SJ, Bailey ST, et al. Diabetic Retinopathy Preferred Practice Pattern®. Ophthalmology. 2025;132(4):P75–P162. doi:10.1016/j.ophtha.2024.12.020

4. Bressler SB, Odia I, Maguire MG, et al. Factors Associated With Visual Acuity and Central Subfield Thickness Changes When Treating Diabetic Macular Edema With Anti–Vascular Endothelial Growth Factor Therapy: An Exploratory Analysis of the Protocol T Randomized Clinical Trial. JAMA Ophthalmol. 2019;137(4):382–390. doi:10.1001/jamaophthalmol.2018.6786

5. Han YE, Jo J, Kim YJ, Lee J. Factors Affecting Intensive Aflibercept Treatment Response in Diabetic Macular Edema: A Real-World Study. J Diabetes Res. 2023;2023(1):1485059. doi:10.1155/2023/1485059

6. Garber AJ. Long-acting glucagon-like peptide 1 receptor agonists: a review of their efficacy and tolerability. Diabetes Care. 2011;34 Suppl 2(Suppl 2):S279–284. doi:10.2337/dc11-s231

7. Vahratian A, Ph.D., M.P.H., Warren A, M.S. Products - Data Briefs - Number 537 - July 2025. September 26, 2025. doi:10.15620/cdc/174616

8. Marso SP, Bain SC, Consoli A, et al. Semaglutide and Cardiovascular Outcomes in Patients with Type 2 Diabetes. N Engl J Med. 2016;375(19):1834–1844. doi:10.1056/NEJMoa1607141

9. Ntentakis DP, Correa VSMC, Ntentaki AM, et al. Effects of newer-generation anti-diabetics on diabetic retinopathy: a critical review. Graefes Arch Clin Exp Ophthalmol. 2024;262(3):717–752. doi:10.1007/s00417-023-06236-5

10. Muayad J, Loya A, Hussain ZS, et al. Influence of Common Medications on Diabetic Macular Edema in Type 2 Diabetes Mellitus. Ophthalmol Retina. 2025;9(6):505–514. doi:10.1016/j.oret.2024.12.006

11. Talebi R, Fortes BH, Yu F, Coleman AL, Tsui I. REAL-WORLD ASSOCIATIONS BETWEEN GLUCAGON-LIKE PEPTIDE-1 RECEPTOR AGONIST USE AND DIABETIC RETINOPATHY ACCOUNTING FOR LONGITUDINAL GLYCEMIC CONTROL. RETINA. 2025;45(9):1663. doi:10.1097/IAE.0000000000004507

12. Phu A, Banghart M, Bahrainian M, Liu TYA, Wolf RM, Channa R. Dipeptidyl peptidase 4 inhibitors, sodium glucose cotransporter 2 inhibitors, and glucagon-like peptide 1 receptor agonists do not worsen diabetic macular edema. J Diabetes Complications. 2024;38(8):108808. doi:10.1016/j.jdiacomp.2024.108808

13. Albert SG, Wood EM, Ahir V. Glucagon-like peptide 1-receptor agonists and A1c: Good for the heart but less so for the eyes? Diabetes Metab Syndr. 2023;17(1):102696. doi:10.1016/j.dsx.2022.102696

14. Tauqeer Z, Bracha P, Hua P, Yu Y, Cui QN, VanderBeek BL. Glucagon-Like Peptide-1 Receptor Agonists are Not Associated with an Increased Risk of Progressing to Vision-Threatening Diabetic Retinopathy. Ophthalmic Epidemiol. 2025;32(4):390–393. doi:10.1080/09286586.2024.2399764

15. Bethel MA, Diaz R, Castellana N, Bhattacharya I, Gerstein HC, Lakshmanan MC. HbA1c Change and Diabetic Retinopathy During GLP-1 Receptor Agonist Cardiovascular Outcome Trials: A Meta-analysis and Meta-regression. Diabetes Care. 2021;44(1):290–296. doi:10.2337/dc20-1815

16. Michaeli T, Khateb S, Levy J. The Effect of Glucagon-like-Peptide-1 Receptor Agonists on Diabetic Retinopathy Progression, Central Subfield Thickness, and Response to Intravitreal Injections. J Clin Med. 2024;13(20):6269. doi:10.3390/jcm13206269

17. Kapoor I, Sarvepalli SM, D’Alessio DA, Hadziahmetovic M. Impact of glucagon-like peptide-1 receptor agonists on diabetic retinopathy: A meta-analysis of clinical studies emphasising retinal changes as a primary outcome. Clin Experiment Ophthalmol. 2025;53(1):67–75. doi:10.1111/ceo.14445

18. Dicembrini I, Nreu B, Scatena A, et al. Microvascular effects of glucagon-like peptide-1 receptor agonists in type 2 diabetes: a meta-analysis of randomized controlled trials. Acta Diabetol. 2017;54(10):933–941. doi:10.1007/s00592-017-1031-9

19. Eleftheriadou A, Riley D, Zhao SS, et al. Risk of diabetic retinopathy and diabetic macular oedema with sodium-glucose cotransporter 2 inhibitors and glucagon-like peptide 1 receptor agonists in type 2 diabetes: a real-world data study from a global federated database. Diabetologia. 2024;67(7):1271–1282. doi:10.1007/s00125-024-06132-5

20. Wai KM, Mishra K, Koo E, et al. Impact of GLP-1 Agonists and SGLT-2 Inhibitors on Diabetic Retinopathy Progression: An Aggregated Electronic Health Record Data Study. Am J Ophthalmol. 2024;265:39–47. doi:10.1016/j.ajo.2024.04.010

21. Marso SP, Bain SC, Consoli A, et al. Semaglutide and Cardiovascular Outcomes in Patients with Type 2 Diabetes. N Engl J Med. 2016;375(19):1834–1844. doi:10.1056/NEJMoa1607141

22. Rubin DB. Bias Reduction Using Mahalanobis-Metric Matching. Biometrics. 1980;36(2):293–298. doi:10.2307/2529981

23. Baker A, Callaway B, Cunningham S, Goodman-Bacon A, Sant’Anna PHC. Difference-in-Differences Designs: A Practitioner’s Guide. J Econ Lit. 2026;64(2):498–557. doi:10.1257/jel.20251650

24. Muayad J, Loya A, Hussain ZS, et al. Influence of Common Medications on Diabetic Macular Edema in Type 2 Diabetes Mellitus. Ophthalmol Retina. 2025;9(6):505–514. doi:10.1016/j.oret.2024.12.006

25. Talebi R, Fortes BH, Yu F, Coleman AL, Tsui I. REAL-WORLD ASSOCIATIONS BETWEEN GLUCAGON-LIKE PEPTIDE-1 RECEPTOR AGONIST USE AND DIABETIC RETINOPATHY ACCOUNTING FOR LONGITUDINAL GLYCEMIC CONTROL. RETINA. 2025;45(9):1663. doi:10.1097/IAE.0000000000004507

26. Ramsey DJ, Makwana B, Dani SS, et al. GLP-1 Receptor Agonists and Sight-Threatening Ophthalmic Complications in Patients With Type 2 Diabetes. JAMA Netw Open. 2025;8(8):e2526321. doi:10.1001/jamanetworkopen.2025.26321

27. Michaeli T, Khateb S, Levy J. The Effect of Glucagon-like-Peptide-1 Receptor Agonists on Diabetic Retinopathy Progression, Central Subfield Thickness, and Response to Intravitreal Injections. J Clin Med. 2024;13(20):6269. doi:10.3390/jcm13206269

28. Kapoor I, Sarvepalli SM, D’Alessio DA, Hadziahmetovic M. Impact of glucagon-like peptide-1 receptor agonists on diabetic retinopathy: A meta-analysis of clinical studies emphasising retinal changes as a primary outcome. Clin Experiment Ophthalmol. 2025;53(1):67–75. doi:10.1111/ceo.14445

29. Talebi R, Fortes BH, Yu F, Coleman AL, Tsui I. REAL-WORLD ASSOCIATIONS BETWEEN GLUCAGON-LIKE PEPTIDE-1 RECEPTOR AGONIST USE AND DIABETIC RETINOPATHY ACCOUNTING FOR LONGITUDINAL GLYCEMIC CONTROL. Retina. 2025;45(9):1663–1671. doi:10.1097/IAE.0000000000004507

30. Wai KM, Mishra K, Koo E, et al. Impact of GLP-1 Agonists and SGLT-2 Inhibitors on Diabetic Retinopathy Progression: An Aggregated Electronic Health Record Data Study. Am J Ophthalmol. 2024;265:39–47. doi:10.1016/j.ajo.2024.04.010

