## Supplemental Materials for "GLP-1 Receptor Agonist Initiation and Anti-VEGF Treatment Frequency in Diabetic Macular Edema: an IRIS^®^ Registry Cohort Study"

Supplemental Table 1. Codes for Inclusion Criteria

| **Subcategory** | **Condition** | **Timing Relative to Index (DME Diagnosis)** | **Codes Included** |
| --- | --- | --- | --- |
| Age | ≥18 years | Anytime | — |
| Diagnosis | Type 2 diabetes with DME | Anytime | ICD-10: E11.311, E11.321X, E11.331X, E11.341X, E11.351X |
| Procedure | Anti-VEGF injections (ranibizumab, bevacizumab, aflibercept) | ≥4 weeks after DME diagnosis | CPT: J2778, J9035, J0177, J0178 |

Supplemental Table 2. Codes for Exclusion Criteria

| **Subcategory** | **Condition** | **Timing Relative to Index (DME Diagnosis)** | **Codes Included** |
| --- | --- | --- | --- |
| Diagnosis | Advanced nonexudative age-related macular degeneration (geographic atrophy with foveal involvement) | Prior to index | ICD-10: H35.3114–H35.3194 |
| Diagnosis | Exudative (wet) age-related macular degeneration | Prior to index | ICD-10: H35.32XX |
| Diagnosis | Branch retinal vein occlusion (BRVO) and related complications | Anytime | ICD-10: H34.23X, H34.83X |
| Diagnosis | Central retinal vein occlusion (CRVO) and retinal artery occlusion | Anytime | ICD-10: H34.1X, H34.81X |
| Diagnosis | Retinal detachment (all types) | Anytime | ICD-10: H33.00X–H33.23 |
| Diagnosis | Chorioretinal inflammation (including uveitis, panuveitis, cyclitis) | Prior to index | ICD-10: H30.X, H44.11X |
| Diagnosis | Endophthalmitis | Prior to index | ICD-10: H44.0X, H44.19 |
| Diagnosis | Hereditary retinal dystrophies | Anytime | ICD-10: H35.5X |
| Diagnosis | Macular structural abnormalities (pucker, scarring, schisis, cysts) | Prior to index | ICD-10: H35.37X, H31.01X, H33.12X |
| Diagnosis | Vitreomacular adhesion | Prior to index | ICD-10: H43.82X |
| Diagnosis | Cystoid macular edema post-cataract surgery | Prior to index | ICD-10: H59.03X |
| Diagnosis | Vitreous hemorrhage | Within 6 months of index | ICD-10: H43.1X |
| Diagnosis | Type 1 diabetes mellitus | Anytime | ICD-10: E10.X |
| Diagnosis | DME not caused by Type 2 diabetes mellitus (secondary, drug-induced, other specified diabetes) | Anytime | ICD-10: E08.X, E09.X, E13.X |
| Procedure | Retinal detachment repair (complex) | Prior to index | CPT: 67108, 67113 |
| Procedure | Pars plana vitrectomy | Prior to index | CPT: 67036–67043 |
| Procedure | Macular laser photocoagulation | Prior to index | CPT: 67210 |
| Medication | Intravitreal steroids (dexamethasone, triamcinolone, fluocinolone) | Prior to index | CPT: J1100, J3301, J7311–J7313 |
| Medication | GLP-1 receptor agonists (e.g., semaglutide, liraglutide, tirzepatide) | Prior to index | RxCUI codes listed |

Supplemental Table 3. Codes for Clinical Diagnosis Covariates

| **Covariate** | **Code System** | **Code** | **Description** | **Timeframe** |
| --- | --- | --- | --- | --- |
| Acute Pancreatitis | ICD-10 | K85.0 | Idiopathic acute pancreatitis | Prior to DME diagnosis |
| Acute Pancreatitis | ICD-10 | K85.1 | Biliary acute pancreatitis | Prior to DME diagnosis |
| Acute Pancreatitis | ICD-10 | K85.2 | Alcohol-induced acute pancreatitis | Prior to DME diagnosis |
| Acute Pancreatitis | ICD-10 | K85.3 | Drug-induced acute pancreatitis | Prior to DME diagnosis |
| Acute Pancreatitis | ICD-10 | K85.8 | Other acute pancreatitis | Prior to DME diagnosis |
| Acute Pancreatitis | ICD-10 | K85.90 | Acute pancreatitis, unspecified | Prior to DME diagnosis |
| Coronary Artery Disease | ICD-10 | I25.1 | Atherosclerotic heart disease | Prior to DME diagnosis |
| Coronary Artery Disease | ICD-10 | I25.10 | Without angina | Prior to DME diagnosis |
| Coronary Artery Disease | ICD-10 | I25.11 | With angina | Prior to DME diagnosis |
| Chronic Kidney Disease | ICD-10 | N18 | Chronic kidney disease | Prior to DME diagnosis |
| Chronic Kidney Disease | ICD-10 | N18.1–N18.6 | CKD stages 1–5, ESRD | Prior to DME diagnosis |
| Chronic Kidney Disease | ICD-10 | N18.9 | CKD unspecified | Prior to DME diagnosis |
| Chronic Kidney Disease | ICD-10 | E08.22, E09.22, E11.22, E13.22 | Diabetes with CKD | Prior to DME diagnosis |
| Cirrhosis | ICD-10 | K74.3–K74.69 | Cirrhosis subtypes | Prior to DME diagnosis |
| Dyslipidemia | ICD-10 | E78.5 | Hyperlipidemia | Prior to DME diagnosis |
| Hypertension | ICD-10 | I10, I15 | Primary and secondary hypertension | Prior to DME diagnosis |
| Gastroparesis | ICD-10 | K31.84, E10.43, E11.43 | Gastroparesis / diabetic autonomic neuropathy | Prior to DME diagnosis |
| Heart Failure | ICD-10 | I50.x | All heart failure subtypes | Prior to DME diagnosis |
| Liver Disease | ICD-10 | K75.89, K72.90 | Inflammatory liver disease, liver failure | Prior to DME diagnosis |
| MASLD / NAFLD | ICD-10 | K76.0 | Fatty liver disease | Prior to DME diagnosis |
| NASH | ICD-10 | K75.81 | Nonalcoholic steatohepatitis | Prior to DME diagnosis |
| Portal Hypertension | ICD-10 | K76.6 | Portal hypertension | Prior to DME diagnosis |
| Stroke | ICD-10 | I63.x | Cerebral infarction (all subtypes) | Prior to DME diagnosis |
| Proliferative Diabetic Retinopathy | ICD-10 | E11.35 | PDR | Anytime |

Supplemental Table 4. Codes for Medication Covariates

| **Medication Class** | **Drug Name** | **Code System** | **Code** | **Exposure Definition** |
| --- | --- | --- | --- | --- |
| Alpha-Glucosidase Inhibitors | Acarbose | RxNorm | 16681 | ≥2 prescriptions over 2 years |
| Alpha-Glucosidase Inhibitors | Miglitol | RxNorm | 30009 | ≥2 prescriptions over 2 years |
| SGLT2 Inhibitors | Empagliflozin | RxNorm | 1545653 | ≥2 prescriptions over 2 years |
| SGLT2 Inhibitors | Canagliflozin | RxNorm | 1373458 | ≥2 prescriptions over 2 years |
| SGLT2 Inhibitors | Dapagliflozin | RxNorm | 1488564 | ≥2 prescriptions over 2 years |
| GLP-1 Receptor Agonists | Semaglutide | RxNorm | 1991302 | ≥2 prescriptions post-index |
| GLP-1 Receptor Agonists | Liraglutide | RxNorm | 475968 | ≥2 prescriptions post-index |
| DPP-4 Inhibitors | Sitagliptin | RxNorm | 593411 | ≥2 prescriptions over 2 years |
| Sulfonylureas | Glimepiride | RxNorm | 25789 | ≥2 prescriptions over 2 years |
| TZDs | Pioglitazone | RxNorm | 33738 | ≥2 prescriptions over 2 years |
| Metformin | Metformin | RxNorm | 6809 | ≥2 prescriptions over 2 years |
| Insulin | Various formulations | RxNorm | Multiple | Any exposure |
| Statins | Atorvastatin | RxNorm | 83367 | Any exposure |
| Statins | Rosuvastatin | RxNorm | 301542 | Any exposure |
| PCSK9 Inhibitors | Evolocumab | RxNorm | 1665684 | Any exposure |
| Antihypertensives | Lisinopril | RxNorm | 29046 | Any exposure |
| Antihypertensives | Losartan | RxNorm | 52175 | Any exposure |
| Diuretics | Furosemide | RxNorm | 4603 | Any exposure |

Supplemental Table 5. Sociodemographic Covariates

| **Covariate** | **Definition** | **Timeframe** |
| --- | --- | --- |
| Age | Age at index event | At index |
| Sex | Recorded biological sex | Anytime |
| Race | Self-reported race | Anytime |
| Ethnicity | Hispanic vs non-Hispanic | Anytime |
| Smoking Status | Ever vs never smoker | Anytime |
| Median Income | Census-linked income category | Anytime |
| High School Graduation Rate | Census-linked education level | Anytime |
| Urban/Rural Status | Census classification | Anytime |
